# Impact of long-term COVID on workers: a systematic review protocol

**DOI:** 10.1101/2022.03.07.22272051

**Authors:** Camila Bruneli do Prado, Giselly Storch Emerick, Luciana Bicalho Cevolani Pires, Luciane Bresciani Salaroli

## Abstract

**Introduction:** Part of the patients infected by COVID-19 have at least one lasting sequel of the disease and may be framed in the concept of long Covid. These sequelae can compromise the quality of life, increase dependence on other people for personal care, impair the performance of activities of daily living, thus compromising work activities and harming the health of the worker. This protocol aims to critically synthesize the scientific evidence on the effects of Covid-19 among workers and its impact on their health status and professional life.

**Method:** Searches will be performed in MEDLINE via PubMed, EMBASE, Cochrane Library, Web of Science, Scopus, LILACS and Epistemonikos. Included studies will be those that report the prevalence of long-term signs and symptoms in workers and/or the impact on their health status and work performance, which may be associated with Covid-19 infection. Data extraction will be conducted by 3 reviewers independently. For data synthesis, a results report will be carried out, based on the main outcome of this study.

**Discussion:** This review will provide evidence to support health surveillance to help decision makers (i.e. healthcare providers, stakeholders and governments) regarding long-term Covid.

## Introduction

On December 31, 2019, the World Health Organization (WHO) was informed of unknown microbial pneumonia cases associated with Wuhan City, Hubei Province, China [1]. It was a new strain of coronavirus that had not been identified before in humans that soon spread rapidly around the world, leading the WHO to declare a public health emergency of international concern on January 30, 2020 [2] and a pandemic on 11 January 2020. March 2020 [3].

This new coronavirus (SARS-CoV-2), responsible for causing the severe acute respiratory syndrome called COVID-19, [4] which has been the focus of attention for scientists, authorities, public health agencies, government officials and communities around the world [5].

As of mid-January 2022, there were 278 million confirmed cases worldwide, including 5.4 deaths, reported to the WHO due to COVID-19 [6]. It should be noted that the total number of subjects who have already been infected is likely to be higher, due to insufficient testing in many countries, as well as the difficulty in diagnosing mild cases of the disease [7].

The acute clinical features of COVID-19 involve pulmonary and extrapulmonary systemic manifestations, causing heterogeneous symptoms such as fever, dyspnea, cough, myalgia, generalized fatigue, anosmia and dysgeusia in milder cases [8]. Of the infected population, 30% evolve without symptoms, 55% have mild and moderate symptoms of the disease, and approximately 15-20% develop more severe manifestations and require hospitalization [9].

Studies show that patients who survive COVID-19, regardless of the clinical form, in the acute phase of the disease, have had a prolonged stay, both in the ICU and in hospital wards, which can evolve to a complex association of cognitive, psychological and motor symptoms, which has been called “Post-COVID-19 Syndrome” [10].

The nomenclature to be used for this complex association of post-COVID symptoms is not yet consolidated and several synonyms have been seen such as “Post-COVID Syndrome”, “Long covid”, “ Late sequelae of COVID-19” or “post-acute COVID-19” [11]. Long-term Covid is conceptually defined as persistent symptoms and/or late complications that people may develop four or more weeks after being infected with the virus that causes COVID-19 [12].

More than 80% of infected patients have at least one lasting sequelae of COVID-19, which may be framed in the concept of long covid [13], causing a decrease in quality of life, increased dependence on other people for personal care and impaired performance of the activities of the daily life [14].

In this context, it is noteworthy that the consequences of COVID-19 can compromise work activities and harm the health of the worker, causing social and economic impacts, since they need to be away from work for health treatment, generating a shortage of manpower. for these áreas [15]. The delay in returning to work considerably increases the chances of losing a job, which can lead to physical and mental problems. When the absence from work lasts more than 6 months, the chance of readaptation to work in the same role drops to 50% [16].

Work capacity is a dynamic process, in which the worker is able to fulfill the requirements of his tasks, using his physical and mental abilities according to his health conditions [17]. Factors such as lifestyle, social characteristics, work requirements, among others, significantly interfere in this process [16].

That said, the SARS-CoV-2 pandemic creates new challenges for occupational health mainly related to the return to work for those affected by long-term symptoms.

Complaints of muscle weakness, memory and concentration difficulties can develop over time, including after returning to work. When this occurs, there is not always recognition or association with COVID-19 [18].

In view of the above, it is relevant to carry out a synthesis of the scientific evidence on the effects of COVID -19 among workers, its impact on their health status and professional life, in order to provide subsidies for the development of follow-up and care actions with workers’ health.

## Methods and analysis

### Research objectives

The objective of this systematic review is to critically synthesize the scientific evidence on the effects of long-term COVID-19 among workers and its impact on health status and working life.

### Search strategy

The search strategy will be carried out in order to increase methodological transparency and improve the reproducibility of the findings, following the PRISMA checklist. Additionally, using the acronym PECO (Population / Exposure / Comparison / Outcomes) [19], we elaborated the guiding question of this review, to ensure the systematic search of the scientific literature: ‘What scientific evidence is available on the impact of long-term COVID-19 on workers? The protocol was registered in the International Prospective Registry of Systematic Reviews (PROSPERO) in November 2021 (registration ID: CRD42021288120).

The search strategy will be carried out through 07 specialized and general databases, from the beginning until April 30, 2021: Medical Literature Analysis and Retrieval System Online (MEDLINE) via Pubmed, EMBASE (Excerpta Medica dataBASE), Cochrane Library, Web of Science, Scopus, Latin American and Caribbean Literature in Health Sciences (LILACS) and Epistemonikos.

In addition, secondary searches will also be carried out in other sources, such as the World Health Organization (WHO) - Global Literature on Coronavirus Disease,, Google Scholar and medRXiv. The reference section of included studies will be manually searched for additional relevant studies. There will be no restriction on publication date or languages in this systematic review. The search strategy will only understand the key terms according to a pre-established PECO acronym. EndNote bibliographic software will be used to store, organize and manage all references and ensure a systematic and comprehensive search. The selection of studies will be performed by three independent researchers (CBP, GSE and LBCP) using Rayyan software.

First, we will identify the existence of subject-specific header indexes in each database (such as MeSH terms, Emtree terms, and DeCS-Health Science Descriptors) and their synonyms (keywords) and additional terms. Search terms will be combined using the boolean operators ‘AND’ and ‘OR’ [20]. The search strategy combining MeSH terms and keywords that will be used in MEDLINE is described in table 1; it will be adapted to meet the specific syntax requirements of each database.

**Table 1.**
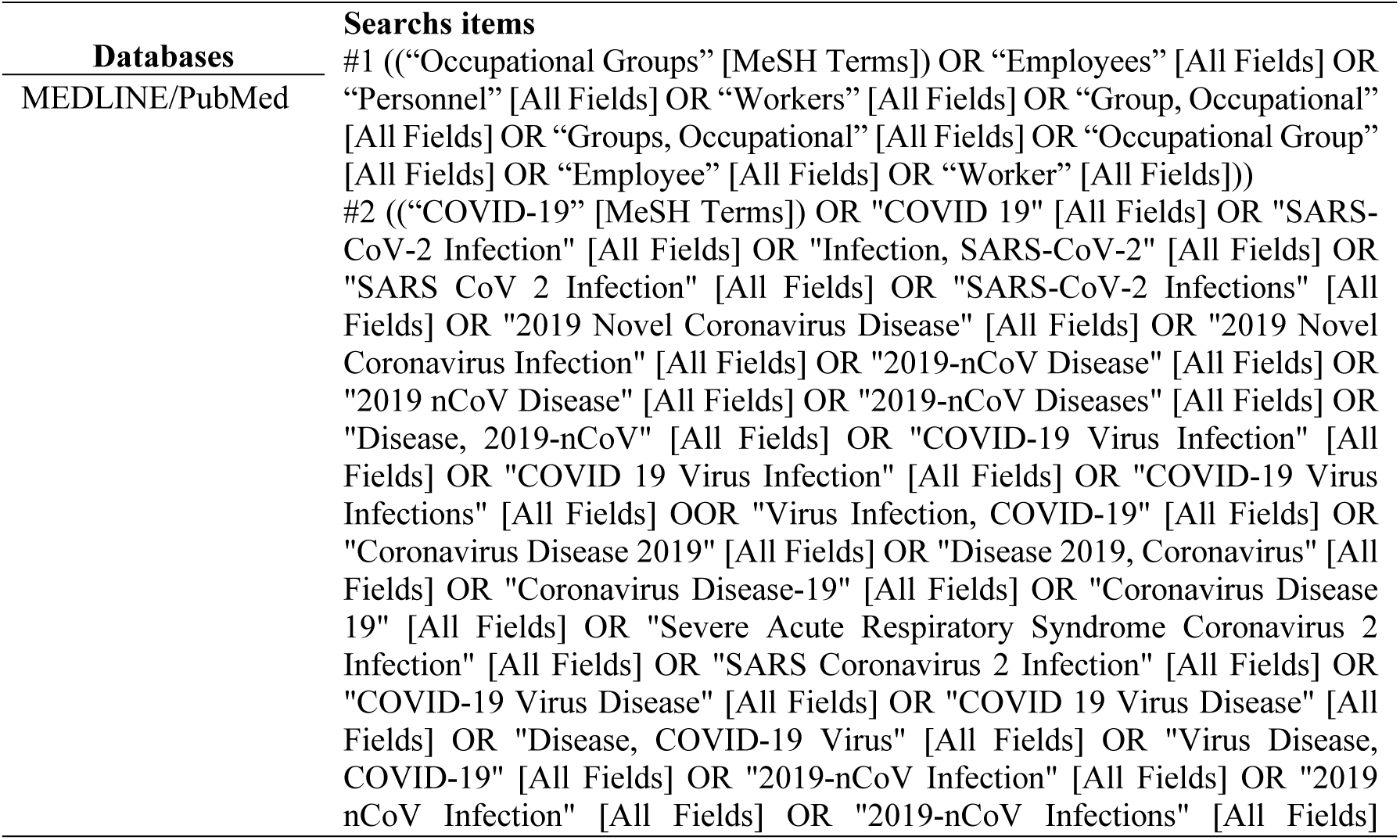
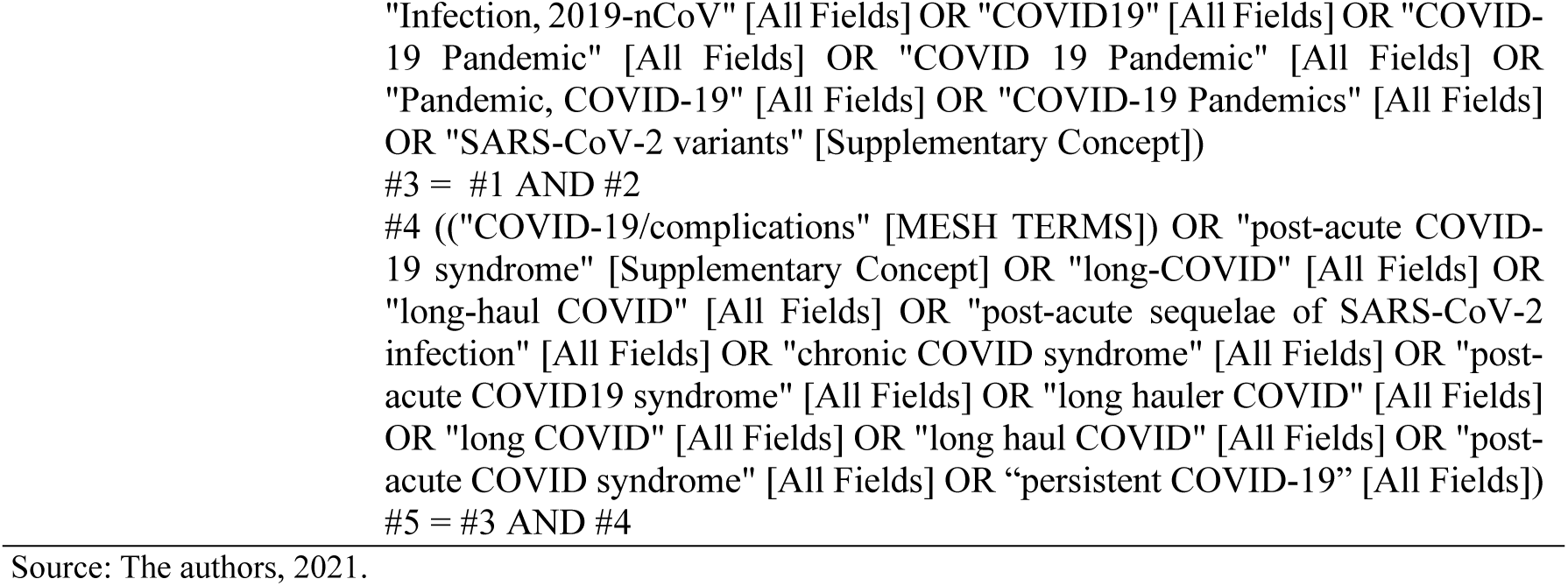
Preliminary pilot Search strategy in MEDLINE/PubMed

| Databases | Searchs items |
| --- | --- |
| MEDLINE/PubMed | #1 ((“Occupational Groups” [MeSH Terms]) OR “Employees” [All Fields] OR “Personnel” [All Fields] OR “Workers” [All Fields] OR “Group, Occupational” [All Fields] OR “Groups, Occupational” [All Fields] OR “Occupational Group” [All Fields] OR “Employee” [All Fields] OR “Worker” [All Fields]))<br>#2 ((“COVID-19” [MeSH Terms]) OR "COVID 19" [All Fields] OR "SARS-CoV-2 Infection" [All Fields] OR "Infection, SARS-CoV-2" [All Fields] OR "SARS CoV 2 Infection" [All Fields] OR "SARS-CoV-2 Infections" [All Fields] OR "2019 Novel Coronavirus Disease" [All Fields] OR "2019 Novel Coronavirus Infection" [All Fields] OR "2019-nCoV Disease" [All Fields] OR "2019 nCoV Disease" [All Fields] OR "2019-nCoV Diseases" [All Fields] OR "Disease, 2019-nCoV" [All Fields] OR "COVID-19 Virus Infection" [All Fields] OR "COVID 19 Virus Infection" [All Fields] OR "COVID-19 Virus Infections" [All Fields] OOR "Virus Infection, COVID-19" [All Fields] OR "Coronavirus Disease 2019" [All Fields] OR "Disease 2019, Coronavirus" [All Fields] OR "Coronavirus Disease-19" [All Fields] OR "Coronavirus Disease 19" [All Fields] OR "Severe Acute Respiratory Syndrome Coronavirus 2 Infection" [All Fields] OR "SARS Coronavirus 2 Infection" [All Fields] OR "COVID-19 Virus Disease" [All Fields] OR "COVID 19 Virus Disease" [All Fields] OR "Disease, COVID-19 Virus" [All Fields] OR "Virus Disease, COVID-19" [All Fields] OR "2019-nCoV Infection" [All Fields] OR "2019 nCoV Infection" [All Fields] OR "2019-nCoV Infections" [All Fields]) |

Possible changes to the protocol will be described in the publication of the final report, which should be developed according to the preferred reporting items for systematic reviews and meta-analyses [21]. The preliminary pilot search strategy combining MeSH terms, synonyms, and keywords to be used in MEDLINE/PubMed is detailed in Table 1.

### Selection of studies

The CEEC strategy is detailed in table 2.

**Table 2.** Inclusion and exclusion criteria.

| PECOS acronym | Inclusion criteria | Exclusion criteria |
| --- | --- | --- |
| P - Patient/Population | Worker of both sexes, of any ethnicity. | Non-workers |
| E - Exposure | Patients with confirmed COVID-19 | Other previous pandemics |
| C - Control or Comparisson | Not applicable | - |
| O - Outcomes | The primary outcomes will be the prevalence of long covid symptoms in workers and/or the impact on their health status and work performance. | ----- |

Source: The authors, 2021.

With regard to study design, primary research studies will be included, including observational studies (e.g. prospective cohort studies, retrospective case reviews), intervention studies (e.g. randomized controlled trials, single-arm studies) and systematic reviews. Secondary research reports (eg non-systematic review articles, news articles, commentary articles, opinion articles) will be excluded. With regard to the characteristics of the population, workers of both sexes and of any ethnicity will be included.

The primary outcomes will be the prevalence of long-term signs and symptoms in workers and/or the impact on their health status and work performance that may be associated with COVID-19 infection.

The screening and selection of studies will be carried out by three reviewers (CBP, GSE and LBCP) independently. Any disagreement will be arbitrated by a fourth reviewer (LBS). The Rayyan application, developed by the Qatar Computing Research Institute, will be used as an auxiliary tool for data management [22].

### Screening

The literature obtained will be imported into Endnote for title and abstract screening, after importing the documents retrieved from the initial searches, duplicates and studies that do not meet the pre-specified inclusion criteria will be excluded and three reviewers (CBP, GSE and LBCP) will independently screen studies based on their titles and abstracts. If there is a good agreement between the reviewers (at least 80%), each one will proceed with the complete screening of the article. If there is agreement below 80%, the articles will be re-evaluated, and differences will be discussed and resolved by consensus; if a disagreement persists, a fourth reviewer (LBS) will make the final decision using the Rayyan app.

After reading the full text of the other studies, the final studies included will be determined. The corresponding author of any original study will be contacted when the full text is not available. The entire study selection process is presented in a PRISMA flowchart [21].

### Data extraction

Full-text screening will be performed by the same independent investigators. To measure agreement between coders during each screening phase, Cohen’s kappa will be performed. Once consensus is reached on the selected studies, a standardized form will be used for data extraction: The information to be extracted includes four domains: (1) study identification (article title, journal title, impact factor, authors, study country, language, funding sources, year of publication, study host institution (hospital, university, research center, single institution and multicenter study), conflicts of interest and study sponsorship); (2) methodological characteristics (study design, study objective or research question or hypothesis, sample characteristics, eg sample size, age, sex, eligibility criteria, ethnicity and baseline characteristics, groups and controls, methods of study recruitment and completion rates, comparator group, time period for follow-up, validated measures, costs and/or fees related to participation, statistical analyzes and adjustments); (3) main findings and implications for clinical practice; and (4) conclusions. The same three reviewers will perform the data extraction independently. Discrepancies between reviewers will be resolved by discussion or, failing agreement, by a fourth reviewer (LBS).

### Methodological evaluation

For internal validity and risk of bias for observational studies, the Methodological Index for Non-Randomized Studies (MINORS) will be used. This MINORS instrument contains eight items for observational studies: (1) a clearly stated objective; (2) inclusion of consecutive patients; (3) prospective data collection; (4) outcomes appropriate to the purpose of the study; (5) unbiased assessment of the study outcome; (6) follow-up period appropriate to the purpose of the study; (7) loss to follow-up of less than 5%; and (8) prospective study size calculation. All items in the MINORS tool will be evaluated from 0 to 2, with a score of 0 indicating that the information was not reported, 1 indicating that the information was reported inappropriately and 2 indicating that the information was reported properly [23].

The same three reviewers (CBP, GSE and LBCP) will independently conduct the quality assessment. Disagreements will be resolved by a fourth reviewer (LBS). Based on the assessment of these domains, studies are classified as low, high, or uncertain risk of bias.

### Data synthesis

If it is not possible to perform meta-analyses, a formal narrative synthesis will be performed according to the Synthesis Without Meta-Analysis (SWiM) guidelines, to obtain data on the prevalence of symptoms of long-term lust in workers and/or the impact on their health status and work performance.

For data synthesis, a results report will be carried out, based on the main outcome of this study. The grouping of studies for synthesis will be based on long and ambiguous outcome, study design (observational studies), risk of bias assessments (studies with low risk of bias), studies that answer the review question. To analyze the quality of the studies, the MINORS tool will be used.

For the critical analysis of the findings, guidelines published by national and international authorities, such as the World Health Organization, the Ministry of Health and the State Health Departments will be used.

To investigate heterogeneity, the data found will be organized into tables, specifying the methodological, sociodemographic characteristics (for example, sex, age), professional category, type of work during the COVID-19 pandemic (remote / face-to-face), severity of symptoms of long covid, clearly referencing all studies included in the review. Prevalence ratio, p-value, confidence intervals, hazard ratio, odds ratio and odds ratio will be described.

Heterogeneity will also be tested by the I^2^ statistic, which can quantify heterogeneity ranging from 0% (no heterogeneity) to 100% (differences between effect sizes can be completely explained by chance alone), and the percentage interpretations are as follows: 0 % -40% indicates potentially unimportant heterogeneity, 30% -60% indicates moderate heterogeneity, 50% -90% indicates substantial heterogeneity, and 75% -100% indicates considerable heterogeneity. To explore heterogeneity across studies, subgroup analysis will be performed using a mixed effects model according to the following variables: age, ethnicity, sex, and occupational characteristics.

### Patient and public involvement

As this is a systematic review protocol, no patient or public will be involved.

### Ethics and disclosure

Due to the characteristics of the design of this study, approval by the ethics committee was not necessary. The results of this systematic review will be disseminated through peer-reviewed publications, as well as through different media such as symposia and conferences related to this field. In addition, any changes to this protocol will be documented with reference to saved searches and analysis methods, which will be recorded in bibliographic databases for data collection and synthesis.

## Discussion

In this systematic review protocol, we clearly describe the study designs, participants, interventions, and outcomes that will be considered according to the research question and data sources, search strategy, data extraction, methodological quality of studies, and synthesis approach. of data. Furthermore, with this protocol study, we reinforced the clarity of the search strategy and minimized the risk of bias. These results will provide evidence to inform and personalize shared decision making for healthcare providers, stakeholders, businesses and government.

Therefore, this systematic review will provide relevant evidence on the effects of long-term COVID among workers and its impact on health status and working life. Ultimately, we will point to evidence in order to provide subsidies for the development of actions to monitor and care for the health of workers and to guide important strategies and decision-makers of health policies in several countries.

To ensure wider dissemination, we will present the interim findings at local and international conferences and publish them in high-impact open access journals.

## Data Availability

N/A

## Acknowledgments

None declared.

